# Highly sensitive and specific detection of the SARS-CoV-2 Delta variant by double-mismatch allele-specific PCR

**DOI:** 10.1101/2021.10.08.21264472

**Authors:** Jeremy A. Garson, Samuel Badru, Eleanor L. Parker, Richard S. Tedder, Myra O. McClure

**Affiliations:** Division of Infection and Immunity, University College London, London, UK; Department of Infectious Disease, St. Mary’s Campus, Imperial College London, London, UK

## Abstract

The highly transmissible Delta variant of SARS-CoV-2 (B.1.617.2), first identified in India, is currently replacing pre-existing variants in Europe, the USA, and many other parts of the world. It is essential to monitor efficiently its spread to help guide public health policies. Genome sequencing is the gold standard for identification of Delta, but is time-consuming, expensive, and unavailable in many regions. We describe here a rapid and relatively inexpensive alternative to sequencing for specific identification of the Delta variant, by application of double-mismatch allele-specific RT-PCR (DMAS-RT-PCR). The technique exploits forward and reverse allele-specific primers, targeting two spike gene mutations, L452R and T478K, within the same amplicon. The discriminatory power of each primer is enhanced by the presence of an additional mismatch located at the fourth nucleotide from the 3′ end. Amplicons are detected in real-time by means of a conventional fluorescently-labelled hydrolysis probe. Specificity was assessed by testing a range of well characterised cell culture-derived viral isolates and clinical samples, most of which had previously been fully sequenced. In all cases the results of viral genotyping by DMAS-RT-PCR were entirely concordant with the results of sequencing, and the assay was shown to discriminate reliably between the Delta variant and other variants of concern (Alpha, B.1.1.7 and Beta, B.1.351), and ‘wild-type’ SARS-CoV-2. Other respiratory viruses, including influenza A and respiratory syncytial virus, were non-reactive in the assay. The sensitivity of DMAS-RT-PCR matched that of the diagnostic SARS-CoV-2 RT-qPCR screening assay, which targets the E gene. Several samples that could not be sequenced due to insufficient virus could successfully be genotyped by DMAS-RT-PCR. The method we describe would be simple to establish in any laboratory that has the ability to conduct PCR assays and should greatly facilitate monitoring of the spread of the Delta variant throughout the world, and its proportional representation in any SARS-CoV-2-infected population.

## Introduction

The global outbreak of COVID-19 which started in China towards the end of 2019 was formally recognised as a pandemic by the World Health Organisation in March 2020. The causative agent, severe acute respiratory syndrome coronavirus 2 (SARS-CoV-2) has been intensively studied since then and more than 3 million genome sequences have been accumulated and made freely available [GISAID, 2021]. As SARS-CoV-2 has evolved, many variants have emerged in different parts of the world and four of these, Alpha, Beta, Gamma and Delta have been designated variants of concern (VOC), primarily on the basis of their increased transmissibility, increased virulence and/or ability to evade immunity. The first three VOC to emerge were the Alpha variant (lineage B.1.1.7), originally identified in the UK, the Beta variant (lineage B.1.351), first identified in South Africa and the Gamma variant (lineage P.1), first identified in Brazil [Davies et al., 2021; Tegally et al., 2021; Faria et al., 2021]. The Delta variant (lineage B.1.617.2) was first detected in India in December 2020 and rapidly increased in prevalence, replacing pre-existing lineages and becoming the most common variant in that country by April 2021. Since then, it has also become the dominant variant in the UK and is out-competing pre-existing variants in Europe, the USA, Israel, and many other countries [Mlcochova et al., 2021; Planas et al., 2021].

The competitive advantage of the Delta variant over other variants is thought to be due to a combination of significantly increased transmissibility and a degree of resistance to both natural and vaccine-induced immunity. There is also evidence that infection with the Delta variant may be associated with higher viral loads, more severe disease, and a greater risk of hospitalisation [Twohig et al., 2021]. It is, therefore, important to monitor the spread of the Delta variant globally in order to inform public health policies and to facilitate timely interventions [Brito et al., 2021]. The gold standard methodology for identifying Delta is full genome sequencing. Although this gives very detailed information and the opportunity to discover new variants, it has the disadvantage of being comparatively expensive, complex, and time-consuming (several days or weeks turnaround time) and is not available in many locations due to lack of expertise and lack of resources. Alternative methods offering rapid, high-throughput, low-cost genotyping for efficient surveillance of the Delta variant are therefore required.

Detection of the S gene by the Applied Biosystems TaqPath Covid-19 PCR assay has been used in the UK as a surrogate marker for the Delta variant [Public Health England, 2021] to differentiate it from the pre-existing Alpha variant, which is characterised by ‘spike gene target failure’ (SGTF) due to the 69-70del S gene mutation. However, this proxy marker method is by no means specific for Delta because both Beta and Gamma variants would also be detected by this strategy [Vogels et al., 2021]. In contrast, the double-mismatch allele-specific RT-PCR (DMAS-RT-PCR) method that we describe here does not exhibit such non-specificity because it targets two separate Delta spike gene mutations, L452R and T478K, within the same amplicon.

The DMAS-RT-PCR technique was first described in 2019 for the genotyping of cancer cell lines [Lefever et al., 2019]. It was subsequently employed for genotyping cacao plant clones [De Wever et al., 2019] but, to our knowledge, has not previously been exploited for genotyping RNA viruses. The very high discriminating power of the technique is achieved by the use of allele-specific primers which, in addition to the 3’ terminal mismatch, have a second artificial mismatch located at the fourth nucleotide from the 3′ end. In the present study, we have validated this approach by designing and evaluating a DMAS-RT-PCR assay that can reliably discriminate between the Delta variant and other SARS-CoV-2 variants. The assay was successfully tested on nose and throat swab samples, and on cell culture-derived SARS-CoV-2 isolates that had previously been characterised by full genome sequencing.

## Materials and methods

### Cell cultured virus samples

Purified SARS-CoV-2 RNA extracted from Vero cell supernatants (2 passages) was kindly provided by Professor Wendy Barclay, Department of Infectious Disease, Imperial College London. RNA was aliquoted and stored at -80°C until use. The following samples were obtained: IC19 (hCoV-19/England/IC19/2020|EPI_ISL_475572|2020-03-17), a B.1 lineage virus with the D614G spike mutation but otherwise the same as the original “wild-type” Wuhan virus. Alpha #246 (hCoV-19/England/205080610/2020|EPI_ISL_723001), an example of the B.1.1.7 variant first identified in the UK. Beta #65 (hCoV-19/England/205280030/2020|EPI_ISL_770441|2020-12-24) and Beta #78 are examples of the B.1.351 variant first identified in South Africa. Delta #395 and Delta #02510 are examples of the B.1.617.2 variant first identified in India. The identities of all the above viruses had been confirmed by full genome sequencing. Influenza A virus PR8 (A/Puerto Rico/8/1934(H1N1) RNA was also provided by Professor Barclay and human respiratory syncytial virus (RSV) strains PP3L and PP3KL were obtained from the laboratory of Dr John Tregoning, Department of Infectious Disease, Imperial College London.

### Clinical samples

Anonymised, residual nose and throat swab samples in virus transport medium were obtained from staff and students at Imperial College London, and from household contacts of individuals with COVID-19 (Research Ethics Committee reference 20/NW/0231, IRAS ID: 282820). Extracted RNA from the samples was stored at -80°C for between 1 day and 8 months before being used in the present study. Initial screening of the samples for SARS-CoV-2 had been performed using a duplex RT-qPCR assay which targets both the E gene (Charité assay) and a human RNA transcript, RNase P (CDC assay) as an internal sample sufficiency control [Rowan et al., 2021].

### Sequencing

In order to establish their viral genotype, clinical samples in which SARS-CoV-2 had been detected were sequenced at the Molecular Diagnostics Unit (MDU), Department of Infectious Disease, Imperial College London, using an Illumina iSeq 100 next-generation sequencing system. In 6 of the samples (see Table 3), the viral concentration was too low for successful sequencing.

**Table 1.**
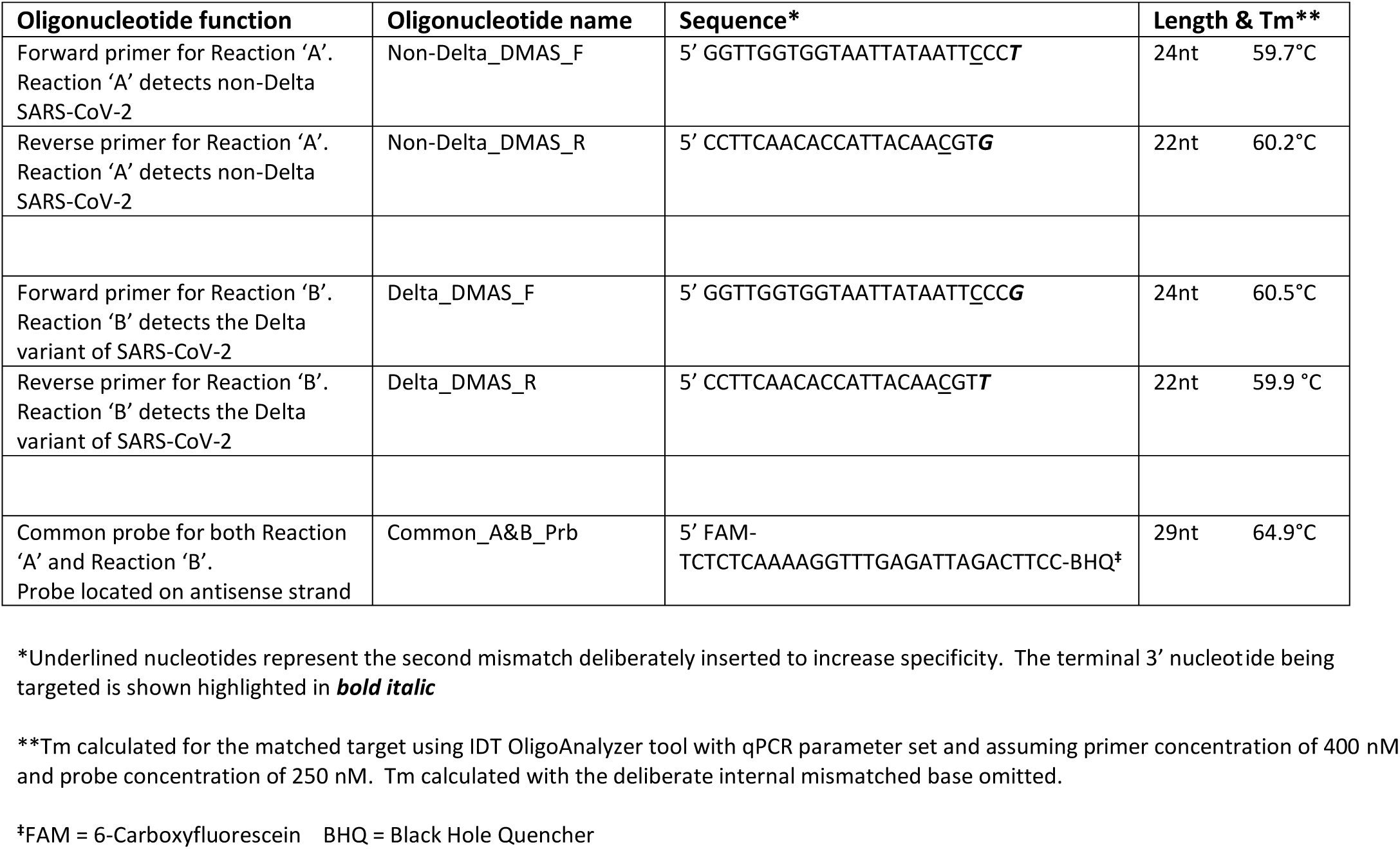
Details of primer and probe sequences used for the DMAS-RT-PCR assay.

**Table 2.** DMAS-RT-PCR findings on cell cultured SARS-CoV-2 isolates.

| Viral Isolate | Genotype by sequencing | Reaction 'A' for non-Delta | Reaction 'B' for Delta | Genotype by DMAS-RT-PCR |
| --- | --- | --- | --- | --- |
| IC19 | 'wild type'* | Detected at all dilutions** | Not detected at any dilution | Not Delta variant |
| Alpha #246 | Alpha variant | Detected at all dilutions** | Not detected at any dilution | Not Delta variant |
| Beta #65 | Beta variant | Detected at all dilutions** | Not detected at any dilution | Not Delta variant |
| Beta #78 | Beta variant | Detected at all dilutions** | Not detected at any dilution | Not Delta variant |
| Delta #395 | Delta variant | Not detected at any dilution** | Detected at all dilutions | Delta variant |
| Delta #02510 | Delta variant | Detected at 1:1,000 only; 17 cycles later than Reaction 'B' | Detected at all dilutions | Delta variant |

\*IC19 is a B.1 lineage virus similar to the original 'wild-type' Wuhan virus but with the D614G spike mutation.
\*\*Ten-fold dilutions 1:1,000, 1:10,000, 1:100,000 and 1:1,000,000 were tested for each isolate.

**Table 3.** Concordance between SARS-CoV-2 genotyping by sequencing and by DMAS-RT-PCR assay.

| Sample No. | Genotype by sequencing | E gene PCR Ct | Reaction 'A' Ct (non-Delta) | Reaction 'B' Ct (Delta) | Genotype by DMAS-RT-PCR | Comments |
| --- | --- | --- | --- | --- | --- | --- |
| 1 | Not done | 19.2 | 37.4 | 20.0 | Delta variant | Expected to be Delta* |
| 2 | Not done | 19.5 | 37.7 | 20.4 | Delta variant | Expected to be Delta* |
| 3 | Not done | 29.6 | Negative | 30.2 | Delta variant | Expected to be Delta* |
| 4 | Not done | 31.1 | Negative | 31.6 | Delta variant | Expected to be Delta* |
| 5 | Not done | 23.6 | Negative | 25.0 | Delta variant | Expected to be Delta* |
| 6 | Not done | 30.1 | Negative | 31.4 | Delta variant | Expected to be Delta* |
| 7 | Not done | 34.0 | Negative | 36.6 | Delta variant | Expected to be Delta* |
| 8 | Insufficient virus | 34.9 | Negative | 34.2 | Delta variant | Expected to be Delta* |
| 9 | Alpha variant | 19.2 | 19.9 | Negative | Not Delta | Non-Delta SARS-CoV-2 |
| 10 | Alpha variant | 27.8 | 27.6 | Negative | Not Delta | Non-Delta SARS-CoV-2 |
| 11 | Alpha variant | 28.3 | 28.6 | Negative | Not Delta | Non-Delta SARS-CoV-2 |
| 12 | Alpha variant | 24.8 | 25.3 | Negative | Not Delta | Non-Delta SARS-CoV-2 |
| 13 | Alpha variant | 26.0 | 25.3 | Negative | Not Delta | Non-Delta SARS-CoV-2 |
| 14 | Alpha variant | 26.5 | 26.4 | Negative | Not Delta | Non-Delta SARS-CoV-2 |
| 15 | Alpha variant | 25.2 | 27.1 | Negative | Not Delta | Non-Delta SARS-CoV-2 |
| 16 | Alpha variant | 23.3 | 25.6 | Negative | Not Delta | Non-Delta SARS-CoV-2 |
| 17 | Alpha variant | 23.8 | 23.7 | Negative | Not Delta | Non-Delta SARS-CoV-2 |
| 18 | Alpha variant | 24.2 | 24.2 | Negative | Not Delta | Non-Delta SARS-CoV-2 |
| 19 | Alpha variant | 35.6 | 33.3 | Negative | Not Delta | Non-Delta SARS-CoV-2 |
| 20 | Alpha variant | 21.7 | 22.0 | 36.3 | Not Delta | Non-Delta SARS-CoV-2 |
| 21 | Alpha variant | 29.3 | 29.6 | Negative | Not Delta | Non-Delta SARS-CoV-2 |
| 22 | Alpha variant | 28.2 | 28.3 | Negative | Not Delta | Non-Delta SARS-CoV-2 |
| 23 | Alpha variant | 27.6 | 28.0 | Negative | Not Delta | Non-Delta SARS-CoV-2 |
| 24 | Alpha variant | 30.6 | 31.1 | Negative | Not Delta | Non-Delta SARS-CoV-2 |
| 25 | Alpha variant | 22.9 | 23.5 | Negative | Not Delta | Non-Delta SARS-CoV-2 |
| 26 | Alpha variant | 24.0 | 25.2 | Negative | Not Delta | Non-Delta SARS-CoV-2 |
| 27 | Delta variant | 30.1 | Negative | 31.2 | Delta variant | Genotypes agree<sup>§</sup> |
| 28 | Delta variant | 23.2 | Negative | 24.2 | Delta variant | Genotypes agree<sup>§</sup> |
| 29 | Insufficient virus | 33.7 | Negative | 34.2 | Delta variant | Expected to be Delta* |
| 30 | Delta variant | 32.0 | Negative | 32.5 | Delta variant | Genotypes agree<sup>§</sup> |
| 31 | Delta variant | 25.4 | Negative | 25.5 | Delta variant | Genotypes agree<sup>§</sup> |
| 32 | Delta variant | 19.0 | Negative | 19.6 | Delta variant | Genotypes agree<sup>§</sup> |
| 33 | Insufficient virus | 34.3 | Negative | 35.5 | Delta variant | Expected to be Delta* |
| 34 | Delta variant | 23.5 | Negative | 24.6 | Delta variant | Genotypes agree<sup>§</sup> |
| 35 | Delta variant | 21.4 | Negative | 21.9 | Delta variant | Genotypes agree<sup>§</sup> |
| 36 | Delta variant | 19.5 | Negative | 20.5 | Delta variant | Genotypes agree<sup>§</sup> |
| 37 | Delta variant | 18.5 | 38.4 | 19.6 | Delta variant | Genotypes agree<sup>§</sup> |
| 38 | Delta variant | 18.1 | Negative | 19.3 | Delta variant | Genotypes agree<sup>§</sup> |
| 39 | Delta variant | 30.7 | Negative | 30.9 | Delta variant | Genotypes agree<sup>§</sup> |
| 40 | Insufficient virus | 35.5 | Negative | 35.3 | Delta variant | Expected to be Delta* |
| 41 | Insufficient virus | 33.9 | Negative | 33.0 | Delta variant | Expected to be Delta* |
| 42 | Insufficient virus | 36.4 | Negative | 38.0 | Delta variant | Expected to be Delta* |

\*Expected to be the Delta variant based on the date of the sample (approximately 99% of samples were Delta variant in UK during July 2021)
<sup>§</sup>Genotypes obtained by sequencing and by DMAS-RT-PCR are in agreement

### RNA extraction

Viral RNA was extracted from clinical samples with a CyBio Felix liquid handing robot (Analytik Jena) and InnuPREP Virus DNA/RNA Kit (Analytik Jena), used according to manufacturer’s instructions. RNA was eluted in 50 µl of RNase Free Water and stored at -80°C until use.

### Primer and probe design

The Delta variant has multiple mutations distributed throughout its genome of which at least five are located within the S gene that encodes the spike protein [Suratekar et al., 2021]. Some of these individual mutations are shared by other variants but we noted that the combination of spike mutations L452R and T478K (corresponding to nucleotide positions T22917G and C22995A, respectively) within the receptor binding domain can effectively differentiate Delta from other variants. Fortuitously, these point mutations are located close enough to each other to permit them to be targeted by a pair of allele-specific primers that generate an amplicon of a length (122 bp) suitable for efficient PCR amplification and detection by a fluorescently-labelled hydrolysis probe. Lefever and co-workers [2019] demonstrated that the discriminatory power of allele-specific primers could be significantly enhanced by adding a second mismatched nucleotide in addition to the mismatch at the 3’ end which targets the mutation. The optimum position for this second internal mismatch was found to be four nucleotides from the 3’ end. We, therefore, designed a pair of double-mismatch allele-specific primers (DMAS) according to these principles. Alignments of SARS-CoV-2 VOC spike gene sequences downloaded from the GISAID database (https://www.gisaid.org/) were performed using MEGA version 7.0.21. The primers (Table 1) were checked by *in silico* PCR (https://genome.ucsc.edu/cgi-bin/hgPcr) to rule out unwanted reactivity with the human genome, and by NCBI BLASTn (https://blast.ncbi.nlm.nih.gov/Blast.cg) to exclude reactivity with other respiratory viruses including human coronaviruses 229E, OC43 and NL63. Melting temperature (Tm) estimations and checks to rule out primer dimers or significant secondary structure were also performed (IDT OligoAnalyzer, https://eu.idtdna.com/calc/analyzer)

### Double-mismatch allele-specific RT-PCR (DMAS-RT-PCR) assay

The method requires two parallel RT-PCR reactions carried out in separate wells. Reaction ‘A’ is designed to detect any SARS-CoV-2 sequence other than the Delta variant, i.e., non-Delta. Reaction ‘B’ is designed to detect only the Delta variant. The primers used for Reaction ‘A’ and Reaction ‘B’ are detailed in Table 1. The amplicons generated in both reactions are detected by a common fluorescently labelled hydrolysis probe (Table 1). Primers and probes were synthesized by Integrated DNA Technologies IDT, Belgium. Five μL RNA template was used in a 20 μL reaction containing 5 μL of 4x TaqMan Fast virus 1-step mastermix (Applied Biosystems), primers at 400 nM and probe at 250 nM. Thermal cycling was performed in a Bio-Rad CFX real-time PCR system with reverse transcription at 54°C for 10 min, followed by 94°C for 3 min, then 40 cycles of 94°C for 15 sec and 58°C for 30 sec. Data were processed using Bio-Rad CFX maestro 2.0 software with baseline subtracted curve fit, fluorescence drift correction, automatically calculated baseline cycles and manual threshold settings. Each run included IC19 RNA and Delta #395 RNA, both diluted a million-fold, as positive controls. Several no-template nuclease-free water negative controls were also included in each run.

### Data Interpretation

The assay is designed primarily for genotyping rather than for quantification and, therefore, does not require a calibration curve. Interpretation of the results is as follows: If in Reaction ‘A’ a sample generates a lower Ct value (i.e., threshold cycle) than in Reaction ‘B’ it indicates that the sample is not the Delta variant. Conversely, if in Reaction ‘B’ a sample generates a lower Ct value than in Reaction ‘A’ it indicates that the sample is the Delta variant. If neither reaction generates a Ct value of less than 40 cycles, this is interpreted as ‘SARS-CoV-2 not detected’.

## Results

### Cell cultured viral isolates

Viral RNA samples extracted from cell culture supernatants were serially diluted in ten-fold steps to cover the range 1:1,000 to 1:1,000,000. The diluted samples were used as templates for Reaction ‘A’ and Reaction ‘B,’ as described under Methods. Fig 1 illustrates that, as predicted, Delta variant RNA (Delta #02510), even at the highest 1:1,000,000 dilution, was efficiently amplified and detected by Reaction ‘B’. In contrast, Reaction ‘A’ was only able to amplify the most concentrated 1:1,000 dilution, but all of the higher dilutions remain undetectable. The 1:1,000 dilution amplification curve crossed the threshold at cycle 39.3 in Reaction ‘A’, almost 19 cycles later than in Reaction ‘B’. The other Delta variant isolate, Delta #395, generated very similar results and was also readily detectable at a 1:1000,000 dilution by Reaction ‘B’ but not detectable at all by Reaction ‘A’, even at the lowest 1:1,000 dilution tested. Conversely, all of the non-Delta viral isolates tested (i.e., IC19, Alpha #246, Beta #65 and Beta #78) were readily detected by Reaction ‘A’ at all dilutions including the highest 1:1000,000 dilution, but not detected at any dilution by Reaction ‘B’. These findings are summarised in Table 2.

**Fig 1.**
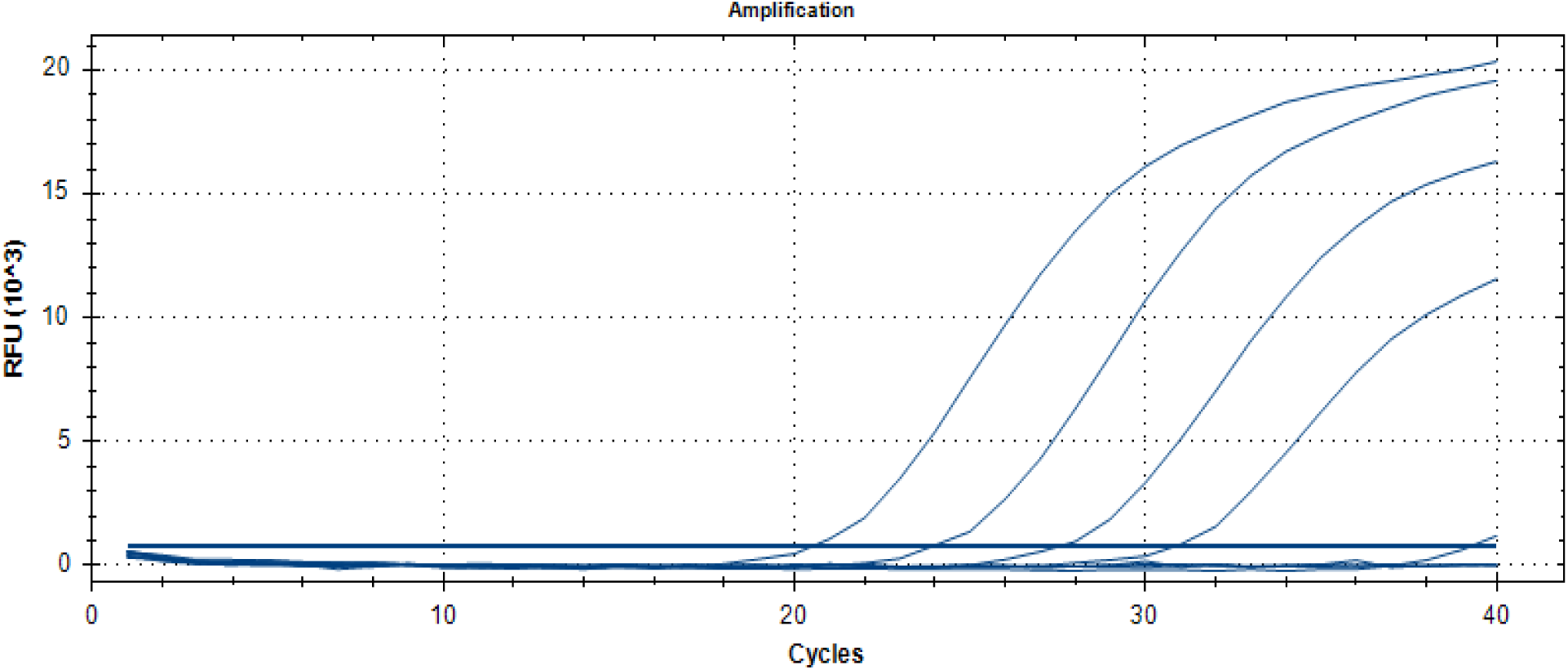
Typical amplification curves generated by Reactions ‘A’ and ‘B’ with dilution series of Delta variant RNA. A ten-fold dilution series of viral isolate Delta #02510 RNA covering the range 1:1,000 to 1:1,000,000 was amplified by Reaction ‘A’ and Reaction ‘B’. Reaction ‘B’ generated the four curves on the left with Ct values of 20.5, 24.0, 27.6 and 30.9. In contrast, Reaction ‘A’ only just detected the lowest 1:1,000 dilution with a Ct value of 39.3 (the very small curve on the extreme right). The three higher dilutions were undetected by Reaction ‘A’.

### Clinical samples

The results generated by testing clinical samples were consistent with those generated by testing cultured viral isolates, in that samples confirmed to be Delta by sequencing, or presumed to be Delta by virtue of the date that they were taken, were efficiently amplified and detected by Reaction ‘B’ but not by Reaction ‘A’. Similarly, samples confirmed non-Delta by sequencing were efficiently amplified and detected by Reaction ‘A’ but not by Reaction ‘B’. The results of testing SARS-CoV-2 positive clinical samples are summarised in Table 3.

Seven of the 42 samples listed in Table 3 (Sample 1 to Sample 7) had not been submitted for sequencing but were expected to be the Delta variant because they were taken from patients during July 2021 when approximately 99% of SARS-CoV-2 infections in the UK were caused by Delta. DMAS-RT-PCR genotyping confirmed that, as expected, all were in fact the Delta variant. Of the remaining 35 samples where sequencing was intended, 6 had such low levels of virus that sequencing was not possible. Sample numbers 8, 29, 33, 40, 41 and 42 all had E gene PCR Ct values greater than 33 cycles. However, all these low virus level samples were successfully genotyped by DMAS-RT-PCR, and all proved to be the Delta variant as expected on the basis of the sample dates.

We observed that four samples (Sample numbers 1, 2, 20 and 37) with very high viral concentrations, corresponding to Ct values ≤ 22 cycles, also produced a weak signal in the non-matching Reaction. For example, Sample 1, a Delta variant with a Reaction ‘B’ Ct value of 20 cycles, generated a very high Ct value of 37.4 in Reaction ‘A’. This is presumed to be due to a very minor degree of non-specific hybridisation of the non-matching primers despite the double mismatch design. This in no way compromises the ability of the DMAS-RT-PCR robustly to distinguish between Delta and non-Delta variants because there is always a very wide gap between the Ct value generated by the two Reactions ‘A’ and ‘B’. Typically, this Ct difference is around 17 cycles (range 14.3 to 18.8 cycles).

There was no evidence of false-positive results being generated by the DMAS-RT-PCR assay. No-template controls were consistently negative and 13 clinical samples that had been negative in the E gene PCR used for SARS-CoV-2 screening were also found to be negative by both Reaction ‘A’ and Reaction ‘B’, i.e., ‘SARS-CoV-2 not detected’.

### Sensitivity

The sensitivity of the DMAS-RT-PCR matched that of the diagnostic SARS-CoV-2 RT-qPCR screening assay which targets the E gene (Charité assay) [Rowan et al., 2021]. All 42 of the clinical samples detected by the E gene PCR screening assay were also detected by the DMAS-RT-PCR and the Ct values generated by the two assays were very similar (Table 3). The mean difference for Alpha variant samples was only 0.3 cycles and the mean difference for Delta variant samples was 0.7 cycles. Rowan et al [2021] demonstrated that the E gene PCR assay has a limit of detection of around 6 SARS-CoV-2 RNA copies per reaction and so we assume that the sensitivity of the DMAS-RT-PCR is approximately the same.

### Specificity

The high specificity of the DMAS-RT-PCR assay enabled it to discriminate reliably between Delta variant and non-Delta variant SARS-CoV-2 with both cell cultured viral isolates and clinical samples. Influenza A and RSV were selected for testing as examples of other common respiratory viruses. The DMAS-RT-PCR did not generate any non-specific signal with either Influenza A virus PR8 or two RSV strains, PP3L and PP3KL.

## Discussion

The DMAS-RT-PCR assay that we describe here for specific identification of the Delta variant differs from previous applications of the technique in that it targets RNA rather than DNA, employs two DMAS primers rather than one, and uses a fluorescently-labelled hydrolysis probe for detection instead of a DNA-binding dye such as SYBR Green. The use of the hydrolysis probe adds an additional layer of specificity to the assay and reduces the chance of false positive signals resulting from primer dimers etc. However, in circumstances where resources are extremely limited, the cost of the assay could be further reduced by replacing the hydrolysis probe with SYBR Green detection. Whether this would have a significant impact on the performance of the assay has yet to be established.

The results presented in this study clearly demonstrate that the Delta variant can be reliably detected and differentiated from other SARS-CoV-2 variants without having to resort to costly, complex, and time-consuming genome sequencing. Resource limitations mean that even in countries where genome sequencing is available it can only be used on a relatively small proportion of positive samples. DMAS-RT-PCR could provide a comparatively inexpensive, simple, rapid, highly sensitive and specific non-commercial alternative to sequencing for epidemiological surveillance of the Delta variant in many settings. An additional advantage of the DMAS-RT-PCR assay is that it appears to be capable of genotyping SARS-CoV-2 when the level of virus in clinical samples is too low for successful sequencing. Counter-intuitively, in countries such as the UK where the Delta variant is already dominant, one might consider using DMAS-RT-PCR for rapidly identifying the small minority of non-Delta cases and subjecting these to genome sequencing in order to increase the chance discovering novel, emergent non-Delta variants. In conclusion, the DMAS-RT-PCR assay that we describe here would be simple to establish in any laboratory that has the ability to conduct PCR assays and should greatly facilitate monitoring of the spread of the SARS-CoV-2 Delta variant globally.

## Data Availability

Full methodological details and summary tabular data are provided in the manuscript.

## Acknowledgments

We are grateful to the NIHR BRC at Imperial College Healthcare Trust for its support of this study. We thank Professor Wendy Barclay and Dr John Tregoning for kindly providing viral isolates used in this study, and Professors Graham Taylor and Ajit Lalvani for their support.

## Author Contributions

Conceptualisation, JAG, RST; Investigation, SB, ELP; Methodology, JAG, SB; Project administration, ELP, RST, MOM; Supervision, JAG, ELP; Writing – original draft, JAG; Writing – review & editing, JAG, SB, RST, MOM; Final approval of submitted version, JAG, SB, ELP, RST, MOM.

